# Patterns and Drivers of Internal Migration and Its Association with Access to Reproductive and Maternity Care Among Women in Pakistan: Evidence from a Recent National Survey

**DOI:** 10.1101/2022.08.04.22278434

**Authors:** Omid Dadras, Takeo Nakayama

## Abstract

**Background:** During the last decades, migratory behavior has had a key role in population growth and redistribution in Pakistan. Migration has far-reaching socioeconomic implications for individuals and society at large that could influence the health integrity of Pakistani women. This study aimed to describe the migration patterns and drivers as well as its association with adequate access to reproductive and maternal care among married Pakistani women aged 15-49.

**Methods:** The data from the 2017-18 Pakistan Demographic Health Survey (PDHS) was used to extract the information on the explanatory (sociodemographic and migration backgrounds) and outcome variables (unmet needs for family planning, adequate antenatal care, and delivery at health facilities). Bivariate and multivariate logistic regression analyses were employed to examine the relationship between these explanatory and outcome variables before and after adjustment for sociodemographic inequalities.

**Results:** In unadjusted models, the odds of having adequate ANC and delivery at health facilities were approximately 2 to 4 times higher in those living (urban non-migrant), moving to (urban to urban, rural to urban), or leaving the urban areas (rural to urban) as compared to rural non-migrants; likewise, the odds of the unmet needs for family planning was about 20-50% lower in the same migration streams compared to rural non-migrant. However, after adjustment for sociodemographic inequalities, most of these associations attenuated and only the association of urban to urban migration with unmet needs for family planning and the association of urban non-migrant with delivery at health facilities remained significant.

**Conclusion:** Although the findings suggest that Internal migration flows, particularly those to urban areas (urban to urban and rural to urban), could be associated with better access to reproductive and maternity care among married Pakistani women aged 15-49 years; adjustment for sociodemographic inequalities, particularly education and wealth, nullified this association to a great extent. This has important implications for current policies and interventions in Pakistan and calls for policy reform and women’s rights advocacy to enhance the literacy level of young Pakistani girls through well-tailored interventions, maintaining them at school.

## Introduction

During the last decades, migratory behavior has had a key role in population growth and redistribution in Pakistan. Migration has far-reaching socioeconomic implications for individuals and society at large (1). Although international migration may have more negative impacts on women than men (2), internal migration has been shown to minimize gender-based socioeconomic inequalities and enhance women’s access to necessary reproductive services and enhance their health integrity (3); however, the knowledge is limited and results are inconsistent across studies (4-6).

In Pakistan, internal migration flows are much larger than international migration flows. Estimates indicate 4 times higher internal migration compared to emigration, equal to approximately 13% of the population moving across cities and districts within the country (7). The most recent estimates of the 2014-2015 Labour Force Survey (LFS) indicated that the main motivations for this movement have been marriage accounting for 35% of total internal migration flows in Pakistan, followed by accompanying the family (21%) and pursuing job opportunities (16.5%). According to the LFS, a quarter of the total internal migration streams belong to rural to urban migration, and men are more likely to migrate as compared to women. Despite this unprecedented urbanization, the national policies are not yet well prepared to address the needs of this huge population; and migrated population, particularly the vulnerable groups such as underprivileged women, may suffer from several financial and human constraints leading to the inability to meet necessary health needs (8, 9).

Several studies have shown that internal migration could empower women through more socioeconomic and educational opportunities in the new environment that had been missing in their previous location and thus, enhance their access to reproductive and maternity care (5, 10-12); while, some studies reported more financial constrain and limited access to necessary reproductive health services among internally migrated women (13, 14). That being said, the ultimate outcome of internal migration among women is more context-specific and not alike across different settings. Against this background, it is important to understand the patterns and drivers as well as the impact that internal migration could have on Pakistani women’s empowerment and their access to necessary reproductive and maternity care. This study aimed to explore the patterns and drivers of internal migration among married Pakistani women aged 15-49 years and explore the impact that it could have on the access to reproductive and maternity care; namely adequate access to family planning, antenatal care (ANC), and delivery at health facilities.

## Method

### Study setting

This study used the data from the Pakistan Demographic Health survey in 2017-18 (PDHS 2017-18) which was the fourth and most recent Demographic and Health Survey conducted in Pakistan. PDHS 2017-18 is a nationally representative population-based survey with the primary objective of providing a comprehensive overview of population, maternal, and child health issues in Pakistan. The survey estimates are also representative for the four provinces of Punjab, Sindh, Khyber Pakhtunkhwa, and Balochistan; for two regions including AJK and Gilgit Baltistan (GB); for Islamabad Capital Territory (ICT); and for FATA. PDHS 2017-18 was conducted under the aegis of the Ministry of Health, Government of Pakistan, with the financial support of the United States Agency for International Development (USAID) and technical assistance from ICF through The DHS Program.

### Sample design

The PDHS 2017-18 employed a stratified two-stage sampling design. The population stratification was accomplished by separating each of the eight regions of the country into urban and rural areas, yielding 16 strata. Samples were selected independently in every stratum through a two-stage selection process. In the first stage, 580 clusters consisting of Ebs (enumeration blocks (EBs) created for the Pakistan Population and Housing Census 2017) were selected by probability proportional to their size, which was the number of households residing in the EB at the time of the census. In the second stage, through an equal probability systematic sampling, a fixed number of 28 households were selected in each cluster which added up to a total of 16,240 households. In the last stage, all the women aged 15-59 years in the household were recruited after informed consent. Due to non-proportional sample allocation, weighting factors were calculated and applied to achieve nationally representative estimates.

### Sample population

The 2017-18 PDHS identified 13,118 ever-married women aged 15-49 who were eligible for interview, of those 12364 women completed the interview, yielding a 94.3% response rate. However; for the purpose of the present study, we only included the currently married women aged 15-49 with a total number of 11,902; because in the PDHS 2018-18, the data for outcome variables were only collected for married women.

### Measurement and scales

Six questionnaires were used to collect the data on key sociodemographic and health indicators in PDHS 2017-18. These include the household questionnaire (HQ), woman’s questionnaire (WQ), man’s questionnaire (MQ), biomarker questionnaire (BQ), fieldworker questionnaire (FWQ), and community questionnaire (CQ) yielding six datasets. The information on patterns and drivers of internal migration was extracted from the HQ data file. The remaining variables related to women’s sociodemography and access to reproductive and maternity care services were extracted for the WQ data file.

### Study variables

#### Outcome variables

We reported three main indicators related to reproductive and maternity care including 1) unmet needs for family planning, 2) adequate antenatal care (ANC), and 3) delivery at health facilities.

##### 1) Unmet needs for family planning

Unmet need was defined as the unmet need for limiting (i.e. women whose most recent pregnancy was not wanted at all, fecund women who did not use contraception despite their desire to have no more children, women who were postpartum amenorrheic for 2 years following an unwanted birth and were not using contraception) and spacing (i.e. women whose most recent pregnancy was not wanted initially but wanted later, fecund women not using contraception who were undecided when/if they wanted a to have a child or who wanted a child 2+ years later, and women who were postpartum amenorrheic for 2 years following a mistimed birth and were not using contraception) (15). The relevant questions had dichotomous response alternatives (i.e., ‘yes’ or ‘no’ responses) and unmet needs for family planning were coded as “yes=1” and “no=0”.

##### 2) Adequate ANC

Based on the World Health Organization (WHO) recommendation, having at least four ANC visits is necessary for optimal maternal and child outcomes [23]. Therefore, adequate ANC was coded as ‘yes=1’ for women with at least four ANC visits before their most recent (4+ ANC visits) in the last five years and coded as ‘no=0’ if there were fewer than four visits.

##### 3) Delivery at health facilities

This variable is coded into “yes=1” indicating delivery at health facilities and “no=0” indicating delivery at home/elsewhere.

### Independent variable

In the PDHS 2017-18, the participants were categorized into six migration streams (urban to urban, urban to rural, rural to urban, rural to rural, urban non-migrant, and rural non-migrant) based on the answer to two questions: “How long have you been living continuously in the current place of residence?” and “Where was the previous place of residence before moving to the current place of residence?”. A further question was collected the data on the drivers of migration from those who migrate either internally or outside the country.

### Covariates

Covariates included in this study were women’s age (15-24, 25-34, and 35-49), education (no education, primary, secondary, higher education), employment (currently working or not), and wealth quintile (poorest, poorer, middle, richer, richest).

### Data analysis

Descriptive statistics were employed to describe the participants’ sociodemographic characteristics and the distribution of migration patterns and drivers (Table 1). Furthermore, the prevalence of the outcome variables was described across migration streams and sociodemographic characteristics, reporting the frequency and percentage for each category of independent variables (Table 2). Bivariate logistic regression was used to examine the likelihood of outcome variables across migratory patterns and sociodemographic factors compared to the reference groups (Table 2), reporting the crude odds ratio (OR) and 95% confidence interval (95% CI). Finally, multivariate logistic regression examines the odds of outcome variables across migration streams compared to the reference group (rural non-migrant), accounting for the effect of sociodemographic factors such as women’s age, education, employment, and wealth index. The results were reported as adjusted odds ratio (AOR) and 95% CI. All the analyses were performed in STATA version 14, taking into account the complex sample design and sampling weight. The significant level was set at a p-value <0.05.

**Table 1.** Distribution of sociodemographic characteristics and reproductive outcomes of currently married women aged 15-49 in PDHS 2017-18

| Variables | N (%) |
| --- | --- |
| **Age group** |  |
| 15-24 | 2490 (20.68) |
| 25-34 | 4781 (40.89) |
| 35-49 | 4631 (38.43) |
| **Education** |  |
| No education | 6405 (48.79) |
| Primary | 1629 (16.64) |
| Secondary | 2232 (21.41) |
| Higher education | 1636 (13.34) |
| **Employment** |  |
| Currently working | 1601 (16.31) |
| Not working | 10299 (83.69) |
| **Wealth index** |  |
| Poorest | 2322 (18.22) |
| Poorer | 2344 (19.42) |
| Middle | 2226 (20.35) |
| Richer | 2342 (20.92) |
| Richest | 2668 (21.10) |
| **Place of living** |  |
| Urban | 5866 (36.77) |
| Rural | 6036 (63.23) |
| **Migration status** |  |
| Migrant | 2467 (19.83) |
| Non-migrant | 9394 (80.17) |
| **Migration stream** |  |
| Urban-Urban | 669 (4.16) |
| Urban-Rural | 320 (3.43) |
| Rural-Rural | 540 (6.97) |
| Rural-Urban | 938 (5.27) |
| Urban non-migrant | 4228 (27.30) |
| Rural non-migrant | 5166 (52.86) |
| **Reason for migration** |  |
| Economic opportunity | 41 (1.8) |
| marriage | 1558 (72.31) |
| accompanied family | 853 (24.64) |
| study | 11 (0.21) |
| transferred on job | 10 (0.23) |
| violence/natural disaster | 15 (0.34) |
| other reasons | 17 (0.43) |
| Total | 11902 (100) |

The sum of numbers may not add up to the total number for some variables due to missing data.

**Table 2.**
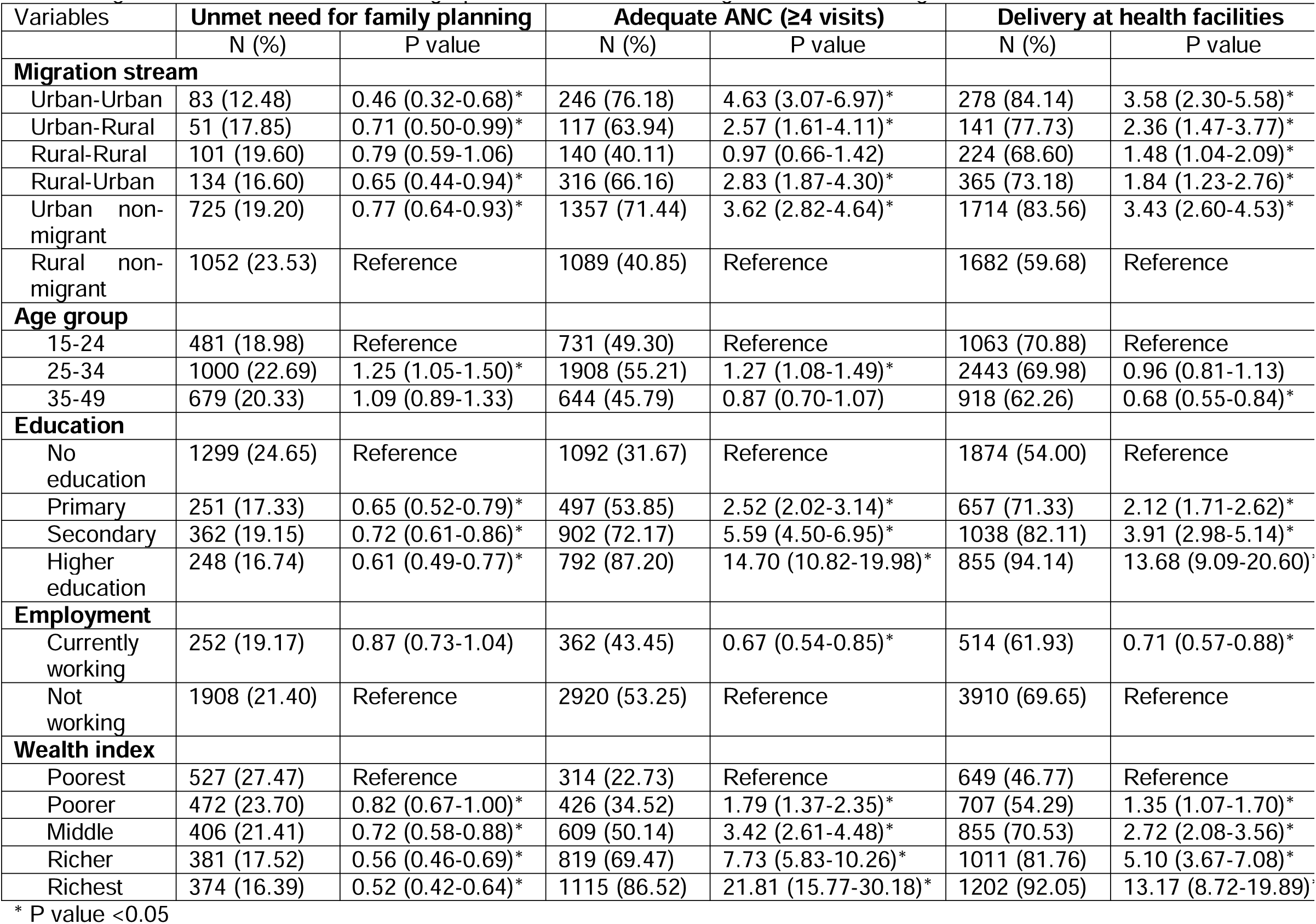
The distribution and likelihood of the unmet need for family planning, adequate ANC, and delivery at health facilities across migration streams and sociodemographic characteristics among married women aged 15-49 in PDHS 2017-18

### Ethical consideration

Data from the PDHS 2017-18 used for this study are available at the DHS website and could be downloaded upon a reasonable request and clear rationale and objectives from the DHS website by a registered user. The ethical approval for conducting the survey was obtained by the ICF International Institutional Review Board (IRB) and the Ethical Review Board of Pakistan Health Research Council before the outset of the survey. An informed verbal and written consent was obtained from all participants prior to the interview. No identity identifier was used in the survey respecting the privacy and confidentiality of participants.

## Result

### 1. The distribution of sociodemographic characteristics of women aged 15-49 in the PDHS 2017-18

As Table 1 indicates, the majority of women were in the 25-34 age group, followed by 35-49, and only a fifth were in the 15-24 age group. Almost half of the women are illiterate and a small proportion (13.34%) has higher education. The majority of women (83.69%) had no job at the time of the interview. There was more or less a fifth of women in each wealth index category. An estimated 63.23% of the participant were urban residents. Almost a fifth were migrants, among whom the rural to rural migrants comprise the majority (6.97%) followed by rural to urban (5.27%). The majority of women mentioned marriage (72.31%) or accompanied family (24.64%) as the main reason for migration.

### 2. The distribution and likelihood of access to reproductive and maternity care across different migration streams and sociodemographic inequalities

The highest unmet need for family planning was observed among rural non-migrant women (23.53%), followed by urban non-migrants and rural to rural migrants. Interestingly the women who moved across rural and urban areas (urban to urban, urban to rural, rural to urban) were less likely to have unmet needs for family planning. The odds of unmet needs were 54%, 29%, 21%, 35%, and 23% lower, respectively, in urban to urban, urban to rural, rural to rural, rural to urban and urban non-migrants as compared to the rural non-migrant women. The prevalence of unmet needs was more or less similar, equal to a fifth of women, across different age groups; however, the likelihood of unmet needs was 25% higher among the 25-34 age group compared to other age groups. As was expected, the women with low literacy were more likely to have unmet needs for family planning and illiterate women had the highest rate of unmet needs (24.65%) followed by primary (17.33%), higher (16.74%), and secondary (19.15%) education groups. There was no significant relationship between the current working status of women and the unmet needs for family planning. It appeared that women of higher wealth have lower unmet needs, and the odds of having unmet needs were 48%, 44%, 28%, and 18% lower, respectively, among the richest, richer, middle, and poorer classes as compared to the poorest class (Table2).

In terms of adequate ANC (≥4), the women who moved across (urban to urban), lived (urban non-migrant), moved to (rural to urban), or left (urban to rural) the urban areas had higher proportions, respectively, 76.18%, 71.44%, 66.16%, and 63,94%, of having adequate ANC as compared to those living in rural areas (40.85%) or moved across rural areas (40.11%). The likelihood of having adequate ANC was approximately 2 to 4 times higher in all migration streams compared to rural non-migrants. It appeared that women aged 25-34 had the highest proportion of adequate ANC compared to other groups and the odds of having adequate ANC was 27% higher among this age group compared to the reference group (15-24). The women with higher education had a substantially higher chance of having adequate ANC; with higher education at the top (OR= 14.70, 95% CI:10.82.19.98) followed by secondary (OR= 5.59, 95%CI: 4.50-6.95), and primary (OR= 2.52, 95%CI: 2.02-3.14). There was a significant association between current working status and having adequate ANC and women currently working were less likely to receive adequate ANC (OR= 0.67, 95%CI: 0.54-0.85). Similar to education, the higher wealth index was associated with a higher number of ANC, and wealthier women were remarkably more likely to have adequate ANC, with the richest class being almost 22 times more likely to have adequate ANC as compared to the poorest class (Table 2). Concerning the delivery in health facilities, we observed relatively higher proportions of health facility delivery among the women who moved across (urban to urban), lived (urban non-migrant), left (urban to rural), and moved to (rural to urban) urban areas as compared to those who were living in rural areas (rural non-migrant) and moved across rural areas (rural to rural) and the odds of health facility delivery was 3.58, 3.43, 2.36, and 1.84 higher, respectively, among urban to urban, urban non-migrant, urban to rural, rural to urban as compared to rural non-migrant women. A lower proportion of health facility delivery was observed in older age groups compared to the youngest and the likelihood of delivery in health facilities was 32% lower in the 35-49 age group as compared to the 15-24 age group. Similar to adequate ANC, higher education and wealth were associated with a higher proportion of health facility delivery and women with the highest education and wealth were almost 13 times more likely to deliver at a health facility as compared to illiterate, and poorest class women (Table 2).

### 3. The impact of internal migration on access to reproductive and maternity care, taking into account the sociodemographic inequalities

Although in the unadjusted model the likelihood of unmet needs for family planning was almost lower in all domains of internal migration as compared to a rural non-migrant stream; after adjusting for sociodemographic inequalities, only urban to urban migrants had a significantly lower likelihood of having unmet needs for family planning (AOR: 0.62, 95%CI: 0.41-0.94) than rural non-migrants.

As Table 3 presents, adjusting for sociodemographic disparities weaken the likelihood of having adequate ANC across all internal migration domains when it was compared to rural non-migrant women in the adjusted model (Table 3).

**Table 3.** Adjusted odds ratio (AOR) and 95% confidence interval (95%CI) for the association of internal migration with unmet need for family planning, adequate ANC, and delivery in health facilities among married women aged 15-49 in PDHS 2017-18

|  | Unmet need for family planning | Adequate ANC (≥4 visits) | Delivery at health facilities |
| --- | --- | --- | --- |
| Migration stream | AOR (95%CI)<sup>1</sup> | AOR (95%CI)<sup>2</sup> | AOR (95%CI)<sup>2</sup> |
| Urban-Urban | 0.62 (0.41-0.94)* | 1.25 (0.81-1.95) | 1.07 (0.67-1.69) |
| Urban-Rural | 0.78 (0.54-1.14) | 1.63 (0.98-2.69) | 1.64 (0.98-2.74) |
| Rural-Rural | 0.84 (0.62-1.14) | 0.70 (0.49-1.01) | 1.21 (0.84-1.75) |
| Rural-Urban | 0.81 (0.54-1.23) | 1.13 (0.73-1.75) | 0.79 (0.53-1.16) |
| Urban non-migrant | 0.98 (0.80-1.21) | 1.25 (0.97-1.60) | 1.37 (1.03-1.83)* |
| Rural non-migrant | Reference<sup>3</sup> | Reference<sup>4</sup> | Reference<sup>4</sup> |

<sup>1</sup> Adjusted for women's age, education, and wealth index.
<sup>2</sup> Adjusted for women's age, education, employment, and wealth index.

The odds of delivery in health facilities were only higher among urban non-migrants (AOR= 1.74, 95%CI: 1.20-2.53) as compared to rural non-migrants in adjuted models (Table 3).

## Discussion

To the best of our knowledge, this is the first study in Pakistan that examined the impact that different migratory patterns could have on the access and utilization of reproductive and maternity care among married Pakistani women aged 15-49 years old. Although the findings indicate only a fifth of the Pakistani women aged 15-49 years are migrants from either rural or urban areas, it appeared that there is a significant relationship between women’s migratory behaviors and access to reproductive and maternity care. The likelihood of having unmet needs for family planning was significantly lower among those who moved from urban to urban, urban to rural, and rural to urban as compared to rural non-migrant women; however, after adjustment for sociodemographic inequalities; only urban to urban migrants were less likely to have unmet needs for family planning. In terms of ANC and place of delivery, those who were living (urban non-migrant) or moved either to (rural to urban) or from (urban to urban/urban to rural) cities were approximately 2 to 4 times more likely to have adequate ANC and deliver at health facilities as compared to rural non-migrant women; however, in adjusted models, only urban non-migrants had a higher likelihood of delivery at health facilities as compared to rural non-migrants. This emphasized the critical role of sociodemographic inequalities particularly the education and wealth that could differ across migratory streams. It is well known that higher education could enhance access to health services regardless of background characteristics (16, 17). This has been attributed to more job opportunities and therefore higher income and welfare as compared to non-educated individuals (18). Similarly in our study, women with higher education and wealth index were more likely to have adequate access to family planning, ANC, and delivery health facilities. Besides, well-educated and economically independent women are more likely to seek necessary reproductive and maternity care through more power to negotiate their reproductive rights with their partner, particularly in man-dominant societies such as Pakistan in which most decisions related to the health of household members are made by men (19, 20).

A study in Peru (21) showed that the utilization of health care services differs among currently married women in urban areas and non-migrant urban women are more likely to seek an institutional source for family planning and ANC as compared with women who are migrants from both urban and rural areas. This has been attributed mainly to the lower level of education of these resettled migrants and could shed light on the attenuated association between utilization of family planning, ANC, and institutional delivery in our study in adjusted models. Moreover; in the Peru study, of three migrant groups (urban non-migrants, urban to urban migrants, and rural to urban migrants), women who were rural migrants were least likely to seek reproductive health services; while, in our study, we observed no significant association between the use of family planning and ANC services among those who move across rural areas (rural to rural) in both unadjusted and adjusted models. On contrary in other studies in Kenya (22, 23) and Myanmar (24), internal migration has been reported as a positive indicator of reproductive and maternity services across all streams of internal migration including rural to rural migration. Although the results of these studies are consistent with our study in unadjusted models, adjusting for sociodemographic inequalities, particularly education and wealth index, produced contradictory results that negate these studies.

Despite these inconsistent findings, one of the most prominent gains that has been acknowledged through internal migration is women empowerment. It has been demonstrated that women from poor households can access opportunities that minimize poverty through migration strategies and lead to more independence, autonomy, and decision-making abilities that could not be achieved otherwise (25). In fact, migration is considered as leverage to freedom from the vicious cycle of poverty among rural women in low-income countries such as Pakistan (26, 27). Women empowerment has been linked to better health integrity and health service utilization (28-30) and we assume the better access to reproductive and maternity care observed among the migrant flows in our study could be medicated by women empowerment moderated by higher education and economic opportunities that migration provides rather than the migration itself. Besides, it is important to understand that although migration may provide women with better opportunities, migrants may not be representative of the total women population in the country as they are prone to positive selection based on wealth or education, which enable them to move (31); however, in our study, it appeared that marriage and accompanying family are the main reasons for migration among Pakistani women which is in line with the findings of previous studies and it seems that this trend has not much changed at least for the last three decades (7, 32). Therefore, the assumption of migration could lead to higher education and economic opportunities at both households and individual levels and consequently higher women empowerment and better access to health service is more arguable. Nonetheless, the first and foremost recommendation should be to empower women by introducing more educational opportunities, maintaining the young girls at school in underprivileged areas in Pakistan through advocacy for women’s rights to higher education in a country with a history of abundance religious and cultural oppression against young women who are seeking independence through education in hope of a better future.

### Limitations

Although this study is one of the first reports that describe the internal migration pattern and its association with access to reproductive and maternity care among married women aged 15-49 years in Pakistan, the findings should be interpreted in light of some limitations. First, it was a cross-sectional study and by nature limited our ability to infer any causal relationship between the outcome variables and explanatory variables as the temporal pattern of the last delivery and antenatal visit against the migration trends was unknown. Second, the data was collected through face-to-face interviews for the last five years on the outcome variables and the last 10 years on the migration patterns; thus, there is a risk of recall bias. Moreover, there is a risk of socially desirable bias for cultural-sensitive questions such as those related to family planning and the use of contraceptives. Another bias that may underestimate the association between internal migration and access to reproductive and maternity care is selection bias that may be introduced if women with higher education and wealth are recruited. Therefore, it is recommended that future surveys recruit a larger number of migrant women to obtain a representative sample of this marginalized group at the national level. Despite all these limitations, the results of this study have important implications for future policy and practices in Pakistan and predict the migratory behavior among Pakistani women and the impact that this movement could have on their access to adequate reproductive and maternity care.

## Conclusion

The findings suggest that Internal migration flows, particularly those to urban areas (urban to urban and rural to urban), could be associated with better access to reproductive and maternity care among married Pakistani women aged 15-49 years; however, this association did not stand after adjustment for sociodemographic inequalities, particularly education and wealth. This emphasizes the important role of higher education and wealth in enhancing access to reproductive and maternity care regardless of the women’s migrant status in both rural and urban areas in Pakistan, Therefore, it is recommended to advocate for women’s right to education to maintain the young girls in the schools and enhance the awareness concerning their reproductive and maternity rights and the necessity of such care for the health integrity of their own and offspring.

## Abbreviations

ANC: Antenatal care
DHS: Demographic Health Survey
USAID: United States Agency for International Development
AOR: Adjusted odds ratio
CI: Confidence interval
OR: Odds ratio

## Declarations

### Consent for publication

Not Applicable

### Availability of data and material

The DHS questionnaire that collected the data in Afghanistan’s demographic and health survey in 2015 could be downloaded from DHS official website (https://www.dhsprogram.com). The dataset (ADHS 2015) that was used in this study could be available upon a reasonable request and with permission from either the Walailak University ethical board or the DHS website.

### Authors’ contribution

OD wrote the research protocol and contributed to the data analysis and writing the manuscript. TN provided critical feedback and comments on the data analysis and results and help in preparing the final draft.

## Acknowledgment

We would like to express our utmost gratitude to the librarians at the library of the University of Bergen for providing technical support to prepare and submit this manuscript.

